# Exercise Stress Echocardiography Unmasks Subclinical Cardiovascular Dysfunction in Midlife Women Following Hypertensive Disorders of Pregnancy

**DOI:** 10.64898/2026.05.21.26353584

**Authors:** Andrea C Kozai, Agnes Koczo, Malamo E Countouris, Tanmay A Gokhale, Takashi Yoshimasu, Benjamin DH Gordon, Janet M Catov

## Abstract

**Background:** Hypertensive disorders of pregnancy (HDP) are a risk factor for early cardiovascular disease in women, perhaps related to adverse cardiovascular reactivity to physiologic stress.

**Objectives:** To evaluate the association of HDP subtypes with exercise stress echocardiography parameters.

**Methods:** This retrospective cohort study linked exercise stress echocardiograms with delivery records. HDP was classified as none, gestational hypertension (GH), and preeclampsia (PEC). We compared features of treadmill exercise stress echocardiography among HDP groups, adjusted for maternal demographic characteristics, time between delivery and stress testing, resting blood pressure (BP), and exercise duration.

**Results:** Among 885 women with matching delivery and exercise echocardiography records (41.4±7.4 years at exercise exam), 92 (10.4%) experienced GH and 39 (4.4%) experienced PEC. Women with PEC were referred for exercise stress testing 3.1 years earlier following delivery (p<0.001) and had shorter exercise duration (ß=-69.6 seconds [95% CI -115.9, -23.4], p=0.003) than those without HDP. Women with GH had higher peak exercise systolic BP (ß=8.96 mmHg [95% CI 4.89, 13.04], p<0.001), diastolic BP (ß=2.67 mmHg [95% CI 0.24, 5.10], p=0.031), and pulse pressure (ß=8.25 mmHg [95% CI 4.11, 12.39], p<0.001) than those without HDP. Women with GH and PEC were twice as likely to have concentric remodeling and more adverse diastolic parameters on echocardiography than those without HDP (p<0.05).

**Conclusions:** Exercise stress echocardiography may detect subclinical cardiovascular dysfunction in midlife women following HDP, with adverse findings differing by subtype: GH was associated with higher peak exercise BP and PEC with lower exercise capacity.

## Introduction

Hypertensive disorders of pregnancy (HDP) are a risk factor for future maternal cardiovascular disease.^1,2^ Women who experience HDP are more likely to develop chronic hypertension within 5-10 years of delivery and are four times more likely to develop heart failure with preserved ejection fraction and other major adverse cardiovascular events than those with normotensive pregnancies.^3^ Adverse structural cardiac remodeling appears prior to the clinical presentation of heart failure, including left ventricular (LV) hypertrophy, concentric remodeling, and evidence of diastolic dysfunction.^4–6^ Similarly, an elevated blood pressure (BP) response to exercise may appear prior to resting criteria for chronic hypertension.^7^ As pregnancy is often described as a “stress test” in revealing future cardiovascular disease risk^1,2^, exercise stress echo may also reveal early manifestations of cardiac maladaptation following HDP prior to clinical presentations of disease.^8,9^

Exercise stress testing is used clinically for patients presenting with symptoms such as chest pain to evaluate for myocardial ischemia, exercise-induced arrhythmias, aerobic fitness, and BP response to exercise.^10^ Poor cardiorespiratory fitness is associated with cardiovascular events and mortality and is an important marker of overt disease risk.^11–13^ In addition, BP response to exercise can provide important insight into cardiovascular reactivity to stress above what can be ascertained from resting BP alone.^10^ Specifically, an exaggerated BP response to maximal exercise is associated with incident hypertension, cardiovascular events, and cardiovascular disease (CVD) mortality.^10,14,15^ Furthermore, women with an exaggerated BP response to exercise are more likely to display abnormal LV remodeling and worse diastolic functional parameters by echo.^10,16,17^ Importantly, cardiorespiratory fitness and BP response to exercise are modifiable with exercise training but may not respond to antihypertensive medications alone, suggesting that exercise prescription may be an important component of CVD risk management particularly in those with poor fitness or abnormal vascular reactivity.^18,19^ In midlife women, exercise testing may be a useful tool to identify those at elevated risk of future clinical cardiovascular sequelae following HDP and inform treatment strategies.

The purpose of this study was to evaluate the differences in exercise capacity, exercise BP responses, and cardiac remodeling among women with versus without a history of HDP who were referred to clinical exercise stress echocardiography. We hypothesized that those who had experienced HDP would have a less favorable exercise stress response, including higher peak BPs, shorter exercise duration, and higher odds of ischemia. Further, we hypothesized those with HDP would demonstrate evidence of unfavorable cardiac adaptations on echocardiography, including higher LV wall thickness and more adverse diastolic parameters.

## Methods

### Study Design and Population

We employed a retrospective cohort design linking electronic health records (EHR) of women who had been referred to and underwent exercise treadmill stress echocardiography within the University of Pittsburgh Medical Center system to delivery records contained in the Magee Obstetric Maternal and Infant (MOMI) Database, a comprehensive record of deliveries at the Magee-Women’s Hospital of University of Pittsburgh Medical Center. This de-identified study was deemed “Exempt” by the University of Pittsburgh Institutional Review Board (STUDY24060017). Inclusion criteria for our cohort specified females who were aged 18-55 at the time of treadmill exercise stress echocardiography and had given birth at least 40 days prior to this evaluation. We restricted our analyses to women without pre-pregnancy hypertension. For women with more than one pregnancy record or more than one encounter of stress echocardiography, only data from the first recorded episode of each were included in the analysis.

### HDP Definition

We used a three-level classification of HDP: 1) no HDP; 2) gestational hypertension (GH); and 3) preeclampsia/eclampsia (PEC). The MOMI Database uses definitions based on those used by the American College of Obstetricians and Gynecologists and abstracted from ICD-9 and -10 codes from the delivery encounter. GH was defined as *de novo* systolic BP ≥ 140 mmHg or diastolic BP ≥ 90 mmHg after 20 weeks’ gestation, according to ICD-9 code 642.3 and ICD-10 code O13. We defined PEC as the *de novo* presence of preeclampsia with or without severe features or eclampsia after 20 weeks’ gestation, including ICD-9 codes 642.4 and 642.5, and ICD-10 code O14. Those classified as No HDP had no diagnosis of hypertension during or before pregnancy. Women with pre-pregnancy hypertension, with or without superimposed PEC (n=43) were excluded from the analysis.

### Exercise Test and Echocardiography Data Abstraction

Echocardiography and exercise stress parameters were ascertained from EHR data. Exercise testing utilized a Bruce protocol in which treadmill speed and grade increased every 3 minutes until volitional fatigue. Heart rate and ischemic changes were monitored using 12-lead electrocardiography. We used reported resting echocardiography values for left ventricular (LV) septal and posterior wall thickness, resting E, A, and e’ velocities, resting and peak exercise systolic and diastolic BP, and time elapsed during the exercise test for analysis. In addition, we calculated 1) relative wall thickness as 2*posterior wall thickness/LV end diastolic dimension, and 2) resting and peak pulse pressure by subtracting diastolic BP from systolic BP. Concentric remodeling was defined as relative wall thickness ≥ 0.42,^20^ and diastolic dysfunction was defined as an average e’ velocity ≤ 6.5 cm/second. Finally, ischemia was defined as ST-segment changes on electrocardiography or wall motion abnormalities on echocardiography during the exercise test, as noted by the interpreting physician.

### Covariates

Time elapsed between delivery and exercise stress echocardiography was ascertained by calculating the number of days between each episode date in the EHR. Maternal age, self-reported race, and insurance status were abstracted from the EHR and included in the MOMI database.

### Statistical Analyses

Participant characteristics were described using means and standard deviations for normally distributed variables, medians and interquartile ranges for skewed variables, and frequencies and percentages for categorical variables. We used multivariable linear or logistic regression to evaluate associations between HDP groups and measures of LV hypertrophy (septal, posterior, and relative wall thickness and presence of concentric remodeling), resting diastolic function parameters (E/A ratio, e’ velocity, E/e’ ratio, and presence of diastolic dysfunction), peak exercise BP response (systolic BP, diastolic BP, and pulse pressure), exercise duration, and presence of exercise-induced ischemia. Given the strong associations between age and exercise parameters^21^, Model 1 was controlled for the time elapsed between delivery and the exercise stress echocardiography exam date for all analyses. A final model (Model 2) included covariate adjustment for maternal age at delivery, race, and insurance status (all analyses), as well as resting BP and exercise time^21^ (BP analyses only). Reported regression estimates are from Model 2. All analyses were conducted using Stata version 18 (StataCorp LLC, College Station, TX). A two-sided significance level of 0.05 was used.

## Results

A total of 9,298 women aged 18-55 underwent 9,808 treadmill exercise stress echocardiography exams between January 2004 and September 2024. Of these, 942 had at least one matching delivery record in the MOMI Database. We excluded one participant with a physiologically improbable change from resting to peak exercise pulse pressure (102 mmHg lower at peak exercise). Our final analytic sample after excluding women with pre-pregnancy hypertension was 885 women (**Figure 1**).

**Figure 1.**
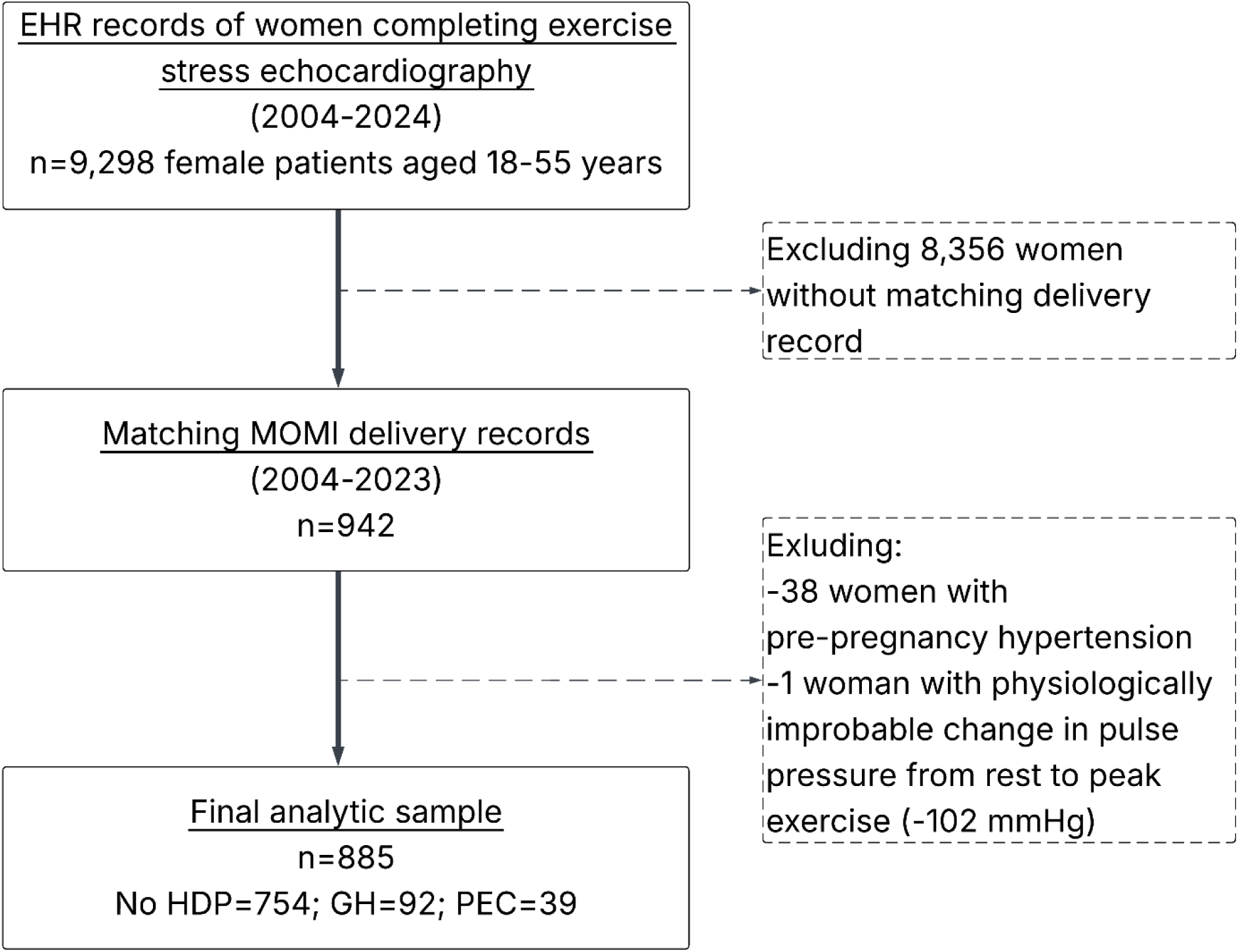
Study flow diagram for the analytic sample. EHR: electronic health record; GH: gestational hypertension; HDP: hypertensive disorders of pregnancy; MOMI: Magee Obstetric Maternal & Infant database; PEC: preeclampsia.

Participant characteristics are reported in **Table 1**. HDP was diagnosed in the index birth in 131 (14.8%) participants; 92 (10.4%) had GH and 39 (4.4%) had PEC. There was no difference among the groups in maternal age at delivery, but women with PEC were significantly younger at the time of exercise stress echocardiography and were referred for testing earlier than those with GH or no HDP (median years after delivery: PEC=5.9; GH=8.0; no HDP=9.0, p<0.001, **Figure 2**). Women with GH or PEC had significantly higher resting SBP and DBP than those with no HDP, but no differences were found among groups for race, insurance status, reason for referral to exercise stress testing (e.g., for symptoms such as chest pain versus prior to a living organ donation procedure), or resting pulse pressure.

**Figure 2.**
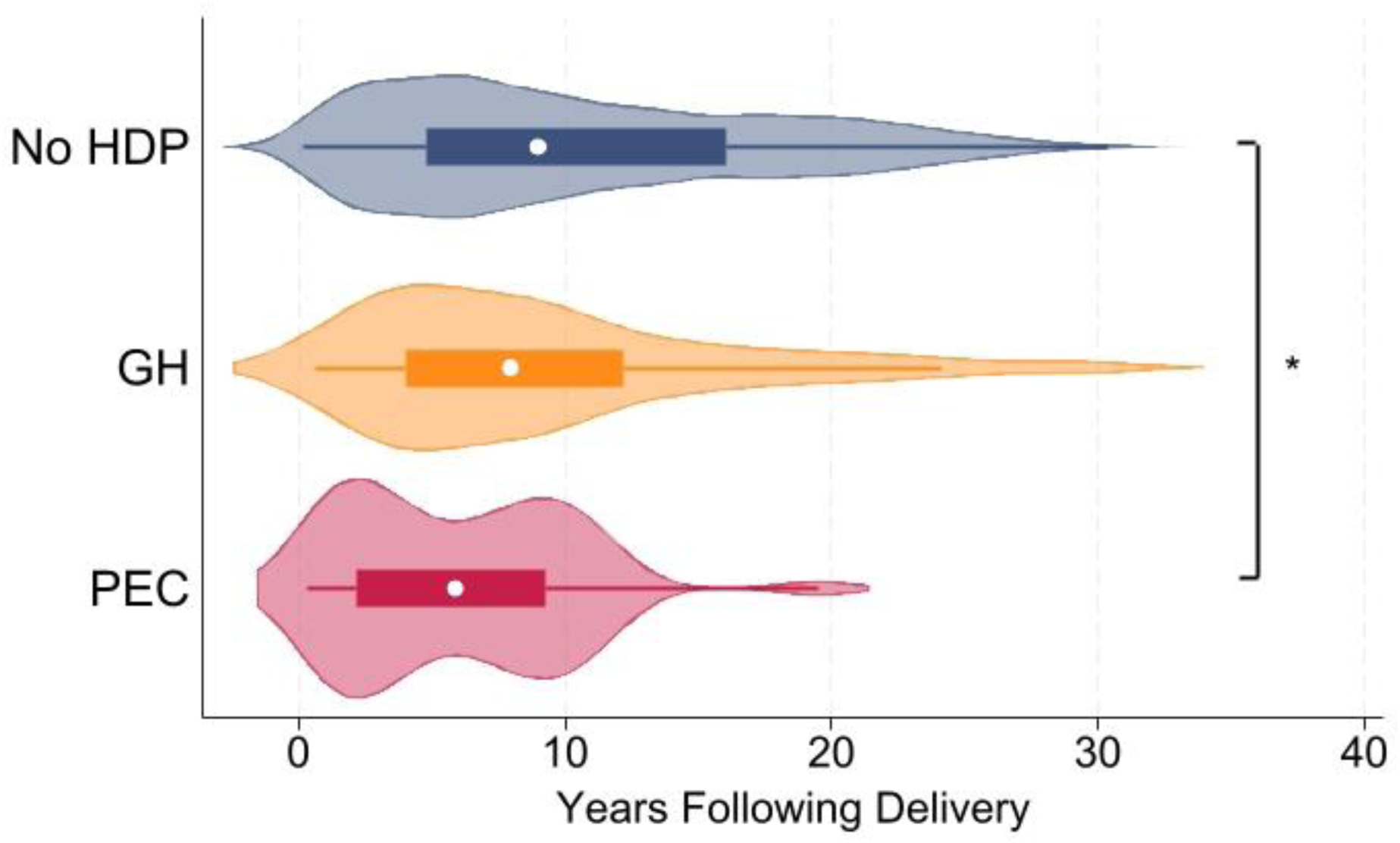
Distribution of time elapsed from delivery to exercise stress echocardiography testing. *indicates p<0.05 for comparison between No HDP and PEC subtypes. GH: gestational hypertension; HDP: hypertensive disorders of pregnancy; PEC: preeclampsia.

**Table 1.**
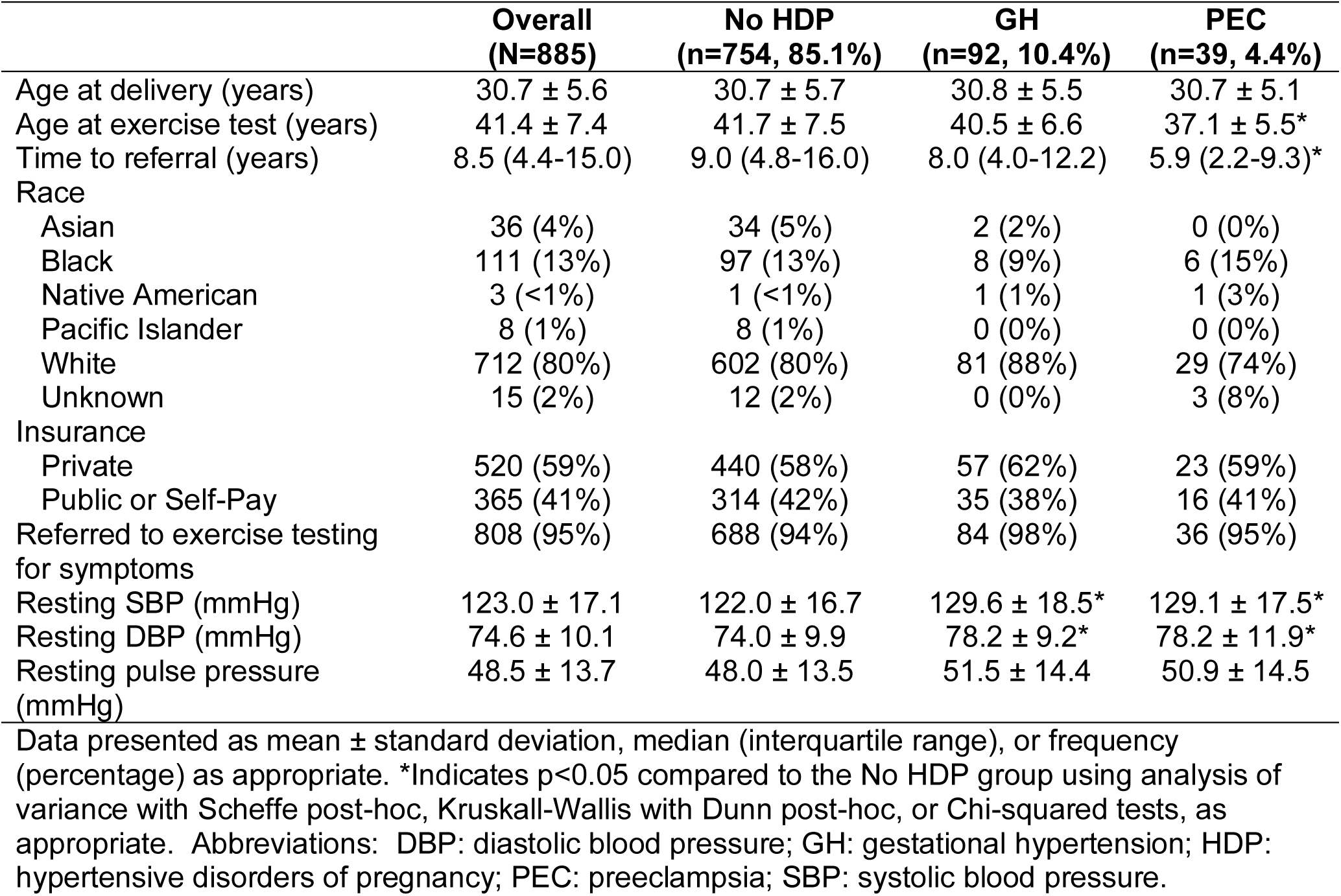
Participant characteristics.

|  | **Overall**<br>**(N=885)** | **No HDP**<br>**(n=754, 85.1%)** | **GH**<br>**(n=92, 10.4%)** | **PEC**<br>**(n=39, 4.4%)** |
| --- | --- | --- | --- | --- |
| Age at delivery (years) | 30.7 ± 5.6 | 30.7 ± 5.7 | 30.8 ± 5.5 | 30.7 ± 5.1 |
| Age at exercise test (years) | 41.4 ± 7.4 | 41.7 ± 7.5 | 40.5 ± 6.6 | 37.1 ± 5.5* |
| Time to referral (years) | 8.5 (4.4-15.0) | 9.0 (4.8-16.0) | 8.0 (4.0-12.2) | 5.9 (2.2-9.3)* |
| Race |  |  |  |  |
| Asian | 36 (4%) | 34 (5%) | 2 (2%) | 0 (0%) |
| Black | 111 (13%) | 97 (13%) | 8 (9%) | 6 (15%) |
| Native American | 3 (<1%) | 1 (<1%) | 1 (1%) | 1 (3%) |
| Pacific Islander | 8 (1%) | 8 (1%) | 0 (0%) | 0 (0%) |
| White | 712 (80%) | 602 (80%) | 81 (88%) | 29 (74%) |
| Unknown | 15 (2%) | 12 (2%) | 0 (0%) | 3 (8%) |
| Insurance |  |  |  |  |
| Private | 520 (59%) | 440 (58%) | 57 (62%) | 23 (59%) |
| Public or Self-Pay | 365 (41%) | 314 (42%) | 35 (38%) | 16 (41%) |
| Referred to exercise testing for symptoms | 808 (95%) | 688 (94%) | 84 (98%) | 36 (95%) |
| Resting SBP (mmHg) | 123.0 ± 17.1 | 122.0 ± 16.7 | 129.6 ± 18.5* | 129.1 ± 17.5* |
| Resting DBP (mmHg) | 74.6 ± 10.1 | 74.0 ± 9.9 | 78.2 ± 9.2* | 78.2 ± 11.9* |
| Resting pulse pressure (mmHg) | 48.5 ± 13.7 | 48.0 ± 13.5 | 51.5 ± 14.4 | 50.9 ± 14.5 |

Data presented as mean ± standard deviation, median (interquartile range), or frequency (percentage) as appropriate. \*Indicates p<0.05 compared to the No HDP group using analysis of variance with Scheffe post-hoc, Kruskal-Wallis with Dunn post-hoc, or Chi-squared tests, as appropriate. Abbreviations: DBP: diastolic blood pressure; GH: gestational hypertension; HDP: hypertensive disorders of pregnancy; PEC: preeclampsia; SBP: systolic blood pressure.

Results of exercise testing analyses are shown in **Table 2**. Women with GH had higher SBP (ß=8.96 mmHg, 95% confidence interval [CI] 4.89, 13.04; p<0.001), higher DBP (ß=2.67 mmHg, 95% CI 0.24, 5.10; p=0.031), and higher pulse pressure (ß=8.25 mmHg, 95% CI 4.11, 12.39; p<0.001) at peak exercise than women with no HDP after adjustment for resting BP, exercise time, time elapsed between delivery and exercise testing, and maternal demographic factors; there were no significant differences in peak BP parameters between women who had PEC versus no HDP. However, women who had PEC had significantly shorter exercise time (ß=-69.6 seconds, 95% CI -115.9, - 23.4) and were significantly more likely to demonstrate ischemia on EKG or echo (odds ratio 6.00, 95% CI 1.77, 20.42) than women with no HDP. Among women with ischemia, detection was by electrocardiography in the majority of cases (No HDP: 68%, GH: 80%, PEC: 100%), with the remainder identified on post-exercise echocardiography.

**Table 2.** Association between HDP status and exercise test parameters.

|  | Mean (SD) | Model 1<br>β coefficient (95% CI) | p-value | Model 2<br>β coefficient (95% CI) | p-value |
| --- | --- | --- | --- | --- | --- |
| **Exercise time (seconds)** |  |  |  |  |  |
| No HDP, n=754 | 563.3 (145.3) | Ref | - | Ref | - |
| GH, n=91 | 540.1 (142.9) | -25.2 (-56.7, 6.3) | 0.117 | -28.4 (-59.2, 2.5) | 0.071 |
| PEC, n=39 | 500.3 (137.0) | **-70.2 (-117.1, -23.2)** | **0.003** | **-69.6 (-115.9, -23.4)** | **0.003** |
| **Peak systolic blood pressure (mmHg)** |  |  |  |  |  |
| No HDP, n=753 | 155.5 (22.1) | Ref | - | Ref | - |
| GH, n=92 | 169.5 (24.0) | **8.72 (4.65, 12.79)** | **<0.001** | **8.96 (4.89, 13.04)** | **<0.001** |
| PEC, n=39 | 160.5 (16.3) | 1.57 (-4.62, 7.77) | 0.618 | 2.35 (-3.90, 8.59) | 0.461 |
| **Peak diastolic blood pressure (mmHg)** |  |  |  |  |  |
| No HDP, n=751 | 77.4 (12.2) | Ref | - | Ref | - |
| GH, n=92 | 82.2 (12.0) | 2.50 (0.08, 4.92) | 0.043 | 2.67 (0.24, 5.10) | 0.031 |
| PEC, n=39 | 78.8 (11.1) | -0.03 (-3.73, 3.67) | 0.987 | 0.29 (-3.46, 4.03) | 0.881 |
| **Peak pulse pressure (mmHg)** |  |  |  |  |  |
| No HDP, n=750 | 78.2 (19.8) | Ref | - | Ref | - |
| GH, n=92 | 87.3 (19.3) | **8.24 (4.10, 12.38)** | **<0.001** | **8.25 (4.11, 12.39)** | **<0.001** |
| PEC, n=39 | 81.7 (19.2) | 3.48 (-2.84, 9.80) | 0.281 | 4.06 (-2.30, 10.43) | 0.210 |
|  | **Frequency (%)** | **Model 1 OR (95% CI)** |  | **Model 2 OR (95% CI)** |  |
| **Exercise-induced ischemia** |  |  |  |  |  |
| No HDP, n=735 | 19 (2.6%) | Ref | - | Ref | - |
| GH, n=91 | 5 (5.5%) | 2.30 (0.83, 6.34) | 0.108 | 2.47 (0.89, 6.90) | 0.084 |
| PEC, n=37 | 4 (10.8%) | **5.55 (1.71, 18.01)** | **0.004** | **6.00 (1.77, 20.42)** | **0.004** |

Model 1 adjusted for time elapsed between delivery and the exercise test, resting blood pressure, and exercise time (blood pressure models only). Model 2 additionally adjusted for maternal age, race, and insurance status. Bold font indicates p<0.05.
Abbreviations: SD: standard deviation; CI: confidence interval; GH: gestational hypertension; HDP: hypertensive disorders of pregnancy; OR: odds ratio; PEC: preeclampsia.

In adjusted analyses of resting echocardiography, women with either GH or PEC had greater LV septal wall thickness(GH: ß=0.07cm, 95% CI 0.03, 0.12; PEC: ß=0.08, 95% CI 0.02, 0.15), posterior wall thickness (GH: ß=0.06cm, 95% CI 0.03, 0.10; PEC: ß= 0.07, 95% 0.02, 0.13), relative wall thickness (GH: ß=0.03, 95% CI 0.01, 0.05; PEC: ß=0.05, 95% CI 0.02, 0.08), and higher odds of concentric remodeling (GH: OR=2.17, 95% CI 1.36, 3.48; PEC: OR=2.08, 95% CI 1.02, 4.24) than those with no HDP (**Table 3**). In addition, e’ velocity was lower in women with GH (ß=-0.74 cm/s, 95% CI -1.46, - 0.01) or PEC (ß=-1.66 cm/s, 95% CI -2.79, -0.53), and the E/A ratio was lower in women with GH compared to no HDP (ß=-0.17, 95% CI -0.29, -0.05), but there was no significant difference in odds of diastolic dysfunction (e’ velocity ≤ 6.5 cm/s) among the groups.

**Table 3.**
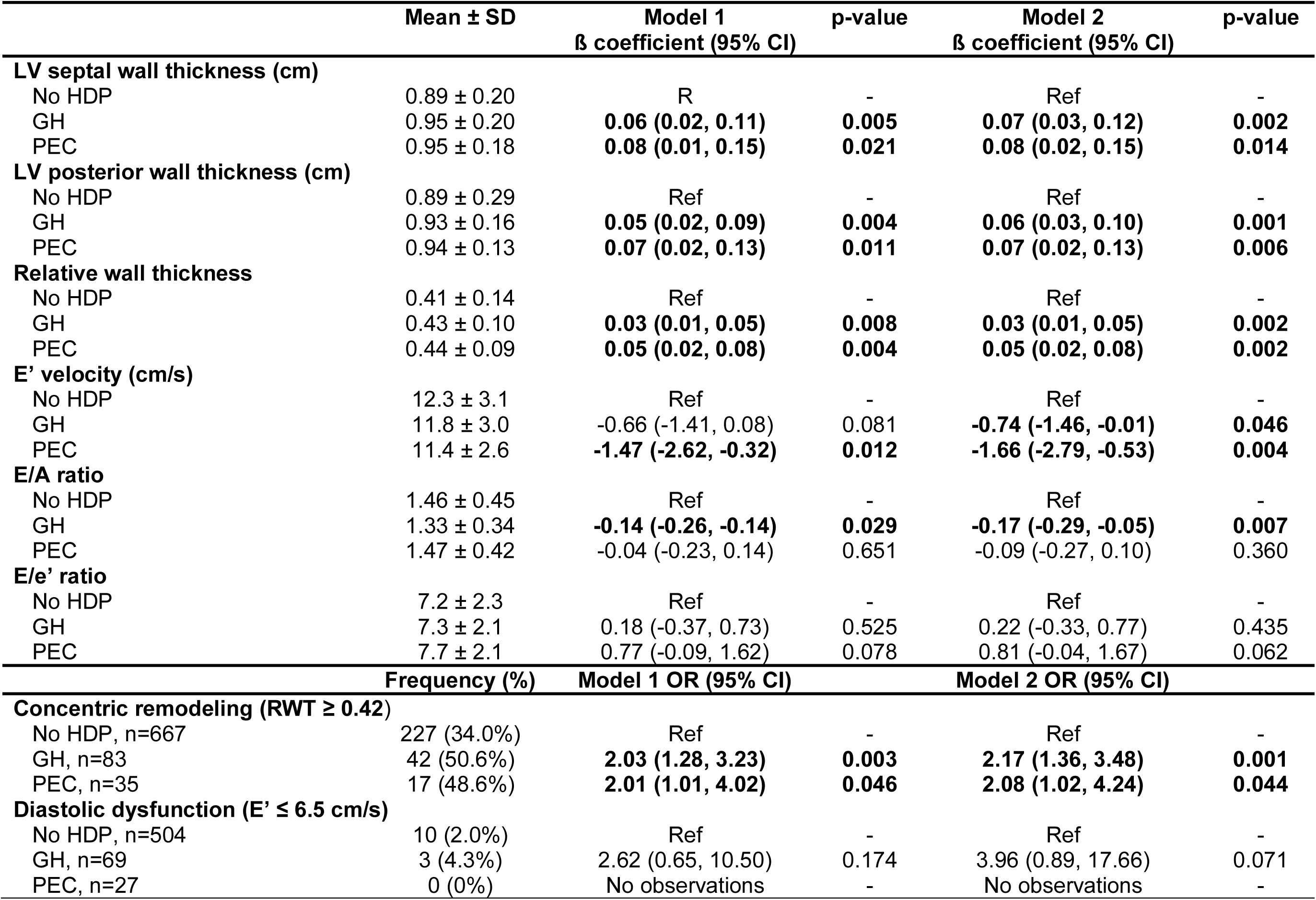
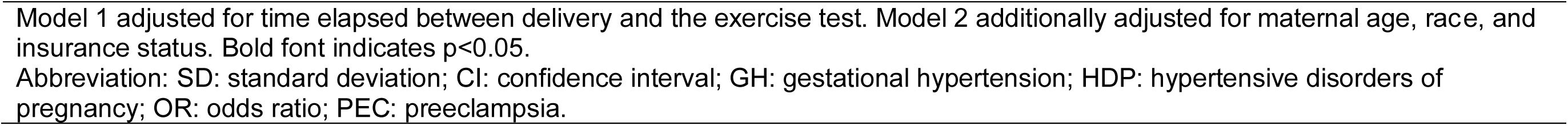
Association between HDP status and resting echocardiography parameters.

|  | Mean ± SD | Model 1<br>β coefficient (95% CI) | p-value | Model 2<br>β coefficient (95% CI) | p-value |
| --- | --- | --- | --- | --- | --- |
| **LV septal wall thickness (cm)** |  |  |  |  |  |
| No HDP | 0.89 ± 0.20 | Ref | - | Ref | - |
| GH | 0.95 ± 0.20 | **0.06 (0.02, 0.11)** | **0.005** | **0.07 (0.03, 0.12)** | **0.002** |
| PEC | 0.95 ± 0.18 | **0.08 (0.01, 0.15)** | **0.021** | **0.08 (0.02, 0.15)** | **0.014** |
| **LV posterior wall thickness (cm)** |  |  |  |  |  |
| No HDP | 0.89 ± 0.29 | Ref | - | Ref | - |
| GH | 0.93 ± 0.16 | **0.05 (0.02, 0.09)** | **0.004** | **0.06 (0.03, 0.10)** | **0.001** |
| PEC | 0.94 ± 0.13 | **0.07 (0.02, 0.13)** | **0.011** | **0.07 (0.02, 0.13)** | **0.006** |
| **Relative wall thickness** |  |  |  |  |  |
| No HDP | 0.41 ± 0.14 | Ref | - | Ref | - |
| GH | 0.43 ± 0.10 | **0.03 (0.01, 0.05)** | **0.008** | **0.03 (0.01, 0.05)** | **0.002** |
| PEC | 0.44 ± 0.09 | **0.05 (0.02, 0.08)** | **0.004** | **0.05 (0.02, 0.08)** | **0.002** |
| **E' velocity (cm/s)** |  |  |  |  |  |
| No HDP | 12.3 ± 3.1 | Ref | - | Ref | - |
| GH | 11.8 ± 3.0 | -0.66 (-1.41, 0.08) | 0.081 | **-0.74 (-1.46, -0.01)** | **0.046** |
| PEC | 11.4 ± 2.6 | **-1.47 (-2.62, -0.32)** | **0.012** | **-1.66 (-2.79, -0.53)** | **0.004** |
| **E/A ratio** |  |  |  |  |  |
| No HDP | 1.46 ± 0.45 | Ref | - | Ref | - |
| GH | 1.33 ± 0.34 | **-0.14 (-0.26, -0.14)** | **0.029** | **-0.17 (-0.29, -0.05)** | **0.007** |
| PEC | 1.47 ± 0.42 | -0.04 (-0.23, 0.14) | 0.651 | -0.09 (-0.27, 0.10) | 0.360 |
| **E/e' ratio** |  |  |  |  |  |
| No HDP | 7.2 ± 2.3 | Ref | - | Ref | - |
| GH | 7.3 ± 2.1 | 0.18 (-0.37, 0.73) | 0.525 | 0.22 (-0.33, 0.77) | 0.435 |
| PEC | 7.7 ± 2.1 | 0.77 (-0.09, 1.62) | 0.078 | 0.81 (-0.04, 1.67) | 0.062 |
|  | **Frequency (%)** | **Model 1 OR (95% CI)** |  | **Model 2 OR (95% CI)** |  |
| **Concentric remodeling (RWT ≥ 0.42)** |  |  |  |  |  |
| No HDP, n=667 | 227 (34.0%) | Ref | - | Ref | - |
| GH, n=83 | 42 (50.6%) | **2.03 (1.28, 3.23)** | **0.003** | **2.17 (1.36, 3.48)** | **0.001** |
| PEC, n=35 | 17 (48.6%) | **2.01 (1.01, 4.02)** | **0.046** | **2.08 (1.02, 4.24)** | **0.044** |
| **Diastolic dysfunction (E' ≤ 6.5 cm/s)** |  |  |  |  |  |
| No HDP, n=504 | 10 (2.0%) | Ref | - | Ref | - |
| GH, n=69 | 3 (4.3%) | 2.62 (0.65, 10.50) | 0.174 | 3.96 (0.89, 17.66) | 0.071 |
| PEC, n=27 | 0 (0%) | No observations | - | No observations | - |

Model 1 adjusted for time elapsed between delivery and the exercise test. Model 2 additionally adjusted for maternal age, race, and insurance status. Bold font indicates $p < 0.05$ .
Abbreviation: SD: standard deviation; CI: confidence interval; GH: gestational hypertension; HDP: hypertensive disorders of pregnancy; OR: odds ratio; PEC: preeclampsia.

## Discussion

In this retrospective cohort analysis of EHR data linking delivery records with subsequent exercise stress echocardiography in midlife women free of prepregnancy hypertension, we found that those with HDP had more adverse responses to exercise stress echocardiography that varied by subtype compared to women with normotensive pregnancy (**Central Figure**). Women who experienced GH had higher peak exercise SBP, DBP, and pulse pressure than those without HDP in the index pregnancy, independent of resting BP. Women who experienced PEC had lower exercise tolerance as determined by the duration of the exercise test, were more likely to demonstrate ischemia, and were referred for clinical exercise stress echocardiography 3 years earlier on average following delivery than women without HDP. Finally, women with either HDP diagnosis had higher LV wall thickness, greater odds of concentric remodeling, and more adverse resting diastolic function parameters than women without HDP.

**Central Figure.**
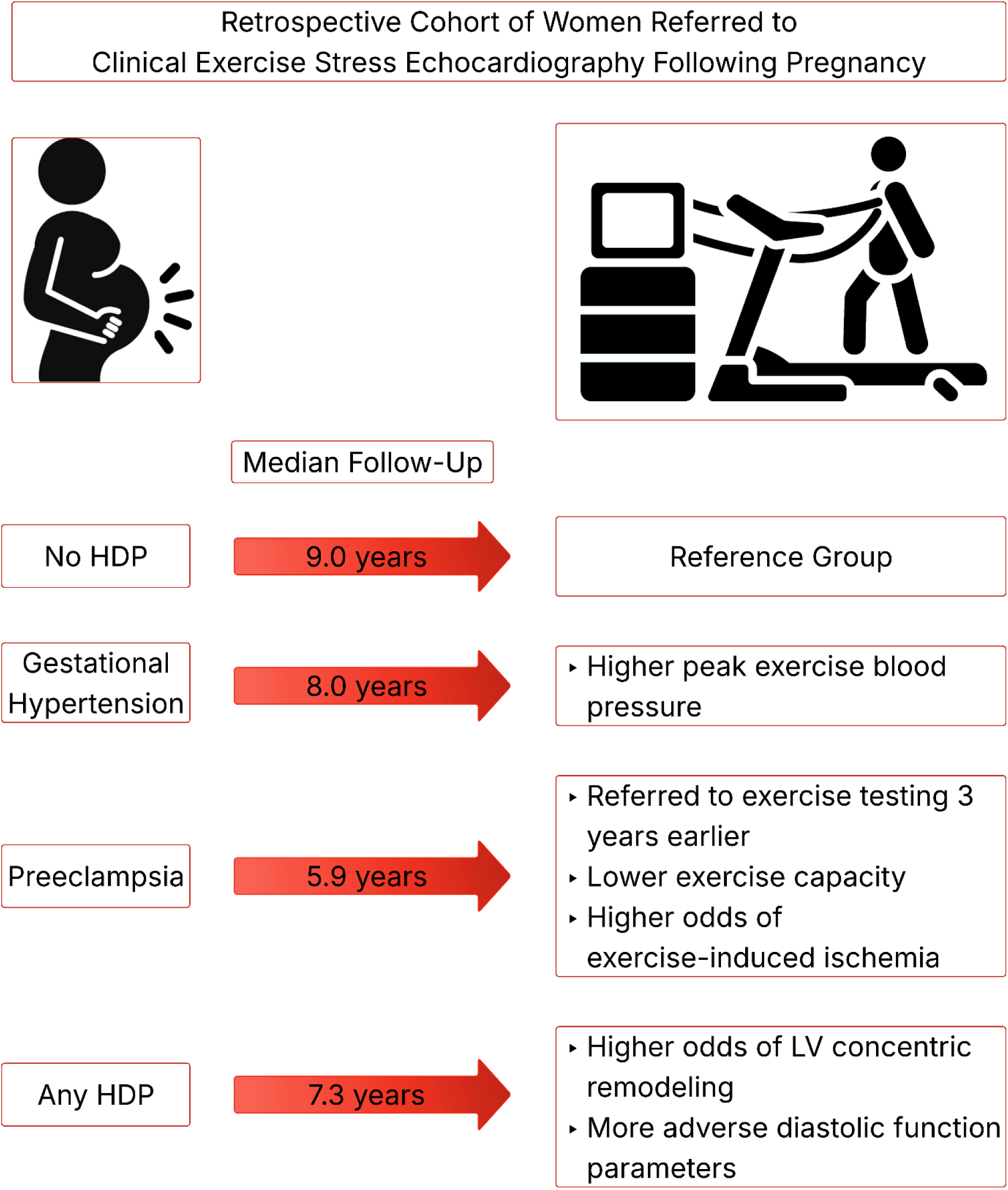
Among women referred to clinical exercise stress echocardiography with a matching birth record, those with a hypertensive disorder of pregnancy (HDP; including gestational hypertension and preeclampsia), demonstrated more adverse exercise and resting echocardiography parameters. LV: left ventricular.

We observed that GH in the index pregnancy was associated with higher SBP, DBP, and pulse pressure at peak exercise. This finding suggests that the functional vascular response to a physiologic stressor is more reactive in these women. These findings were independent of resting BP, suggesting that GH may confer excess vascular reactivity not fully captured by resting measurements.^21^ Exercise BP may reflect the hemodynamic load experienced during physical activity in daily life, implying that the true daily vascular burden in these women extends beyond what resting assessments reveal. Further, we found higher pulse pressure in those who experienced GH. While exercise pulse pressure is less explored in its relationship to CVD risk, widening resting pulse pressure over time is recognized as a marker of vascular aging^22^ and precursor to CVD.^23^ It is possible that our observation of greater pulse pressure at maximal exercise is an early indicator of risk in this midlife sample; this hypothesis should be evaluated with future research.

Women with PEC in the index pregnancy exhibited significantly lower exercise capacity than those with no HDP, despite undergoing testing at younger ages where expected exercise tolerance would be higher. Our observed difference in time to peak exercise was approximately 70 seconds shorter in those with PEC, suggesting a lower VO2max of approximately 1-2 METs compared to those without HDP.^24^ In women, each 1-MET higher exercise capacity has been associated with 23% lower risk of cardiac events^25^, suggesting the potential clinical relevance of this finding as a key prognostic indicator of CVD risk.^26–28^ These data expand upon prior literature demonstrating lower exercise capacity within three months of delivery among women with PEC.^29^ Our results suggest that PEC may be related to poor cardiorespiratory fitness, though we cannot determine from the present study whether poor fitness preceded or resulted from PEC. Prior research in the Coronary Artery Risk Development in Young Adults Study found that cardiorespiratory fitness did not predict the incidence of preterm birth or small-for-gestational-age birth^30^, but deliveries complicated by HDP were not included in the analysis. Regardless of directionality, cardiorespiratory fitness is modifiable with exercise training.^31–33^ Our results suggest that exercise training after PEC should be investigated as a risk-reducing strategy.^34^

In addition to poorer cardiovascular fitness, we observed higher rates of ischemia on exercise testing in women with PEC (10.8% compared to 2.5% in those without HDP). We also observed a higher rate of ischemia in women with GH (5.5%), but it did not reach statistical significance, likely reflecting insufficient power. The small number of ischemic outcomes and subsequently large confidence intervals suggests these results should be interpreted with caution, but prior literature supports the hypothesis that HDP is associated with accelerated development of ischemic heart disease.^3,35^ It is particularly notable that ischemia was present at higher frequencies among those with prior PEC given the relatively young age at which these women were evaluated, emphasizing the importance of close monitoring following a delivery complicated by HDP. PEC may confer a cumulative vascular burden that persists following delivery and puts these women at an elevated risk of early CVD. In support of this hypothesis, prior work by our group found that among women who experience a myocardial infarction before the age of 65, women with a history of PEC reached this endpoint 4.5 years sooner following delivery than those with uncomplicated pregnancies.^35^ Results of our current analysis demonstrate that subclinical markers like elevated exercise BP and poor cardiorespiratory fitness manifest sooner after delivery in those with PEC. Therefore, exercise testing may be a useful tool to identify those at greatest risk following HDP.

On echocardiographic analysis, our study noted several important findings. Structurally, women had progressively greater left ventricular wall thickness and relative wall thickness by HDP severity (no HDP< GH< PEC). Further, we observed double the odds of concentric remodeling in those with an HDP diagnosis compared to those without HDP. Coupled with our finding of higher odds of ischemia, this may indicate myocardial damage related to elevated hypertensive burden or microvascular dysfunction.^36^ Given the greater incidence of heart failure with preserved ejection fraction among midlife women following HDP^1,37–39^, it is possible concentric remodeling contributes to the increase in clinical heart failure presentations. Management strategies to reverse concentric remodeling in this subclinical phase are critically needed to mitigate further cardiovascular morbidity.

Finally, women with HDP had worse measures of diastolic function including lower e’ tissue doppler velocity, lower E/A ratio and higher E/e’ ratio; these findings varied by subtype and were typically most severe among those with PEC. However, we did not detect differences in presence of clinical diastolic dysfunction. Notably, very few women in this study reached diagnostic criteria for diastolic dysfunction, suggesting this parameter may be a later-stage indicator of disease risk. Among women without prepregnancy hypertension being evaluated in the first decade following HDP, exercise parameters and LV remodeling may be more sensitive clinical outcomes.

### Strengths and Limitations

A strength of the study was the use of real-world data in the EHR, which allowed us to generalize our findings to similar clinical samples. Additionally, by matching the health records of women with exercise testing to our robust pregnancy outcomes database, we were able to ensure our HDP exposure was appropriately classified. Furthermore, our analyses were limited to women who were normotensive prior to pregnancy, which limited confounding from pre-existing hypertension and demonstrated the degree of cardiovascular burden seen following *de novo* hypertension in pregnancy. However, certain limitations must also be acknowledged. Submaximal BPs were not recorded in the EHR, which may limit our interpretations due to accuracy of peak BP ascertainment and differing maximal workloads.^21^ Our analyses were adjusted for exercise time to address this limitation. Additionally, our analyses did not account for HDP status in subsequent pregnancies; future research should evaluate exercise outcomes accounting for additional pregnancies. Finally, our sample was restricted to women who were referred to clinical exercise testing, predominately due to symptoms such as chest pain. We are unable to generalize our findings to a healthier general population, but the presence of poorer exercise and echocardiographic parameters in those with HDP even compared to women who developed symptoms without an HDP exposure suggests that HDP is likely a strong predictor of subsequent cardiovascular dysfunction detectable in clinical records in relatively young women.

## Conclusions

In this retrospective EHR cohort study, we found that women who experienced HDP had more adverse exercise stress and resting echocardiography parameters on clinical exercise testing about a decade after delivery. Elevated exercise BPs and higher odds of ischemia and concentric remodeling demonstrate the importance of close monitoring after HDP. Midlife is a crucial time in which clinical disease progression may be detectable using exercise stress echocardiography in women following HDP. Finally, the potential for improved exercise capacity as a cardioprotective strategy for women with HDP warrants further investigation.

## Data Availability

Data were obtained from the Steve N. Caritis MWRI Magee Obstetrical Maternal Infant (MOMI) Database and Biobank supported by the RK Mellon Foundation and the University of Pittsburgh Clinical and Translational Science Institute (5UL1 TR001857-02).

## References

1. Lane-Cordova AD, Khan SS, Grobman WA, Greenland P, Shah SJ. Long-Term Cardiovascular Risks Associated With Adverse Pregnancy Outcomes. Journal of the American College of Cardiology. 2019;73:2106–2116.

2. Khan SS, Cameron NA, Lindley KJ. Pregnancy as an Early Cardiovascular Moment: Peripartum Cardiovascular Health. Circulation Research. 2023;132:1584–1606.

3. Giorgione V, Jansen G, Kitt J, Ghossein-Doha C, Leeson P, Thilaganathan B. Peripartum and Long-Term Maternal Cardiovascular Health After Preeclampsia. Hypertension. 2023;80:231–241.

4. Giorgione V, Khalil A, O’Driscoll J, Thilaganathan B. Peripartum Screening for Postpartum Hypertension in Women With Hypertensive Disorders of Pregnancy. JACC. 2022;80:1465–1476.

5. Countouris ME, Villanueva FS, Berlacher KL, Cavalcante JL, Parks WT, Catov JM. Association of Hypertensive Disorders of Pregnancy With Left Ventricular Remodeling Later in Life. J Am Coll Cardiol. 2021;77:1057–1068.

6. Hauspurg A, Countouris ME, Catov JM. Hypertensive Disorders of Pregnancy and Future Maternal Health: How Can the Evidence Guide Postpartum Management? Curr Hypertens Rep. 2019;21:96.

7. Manolio TA, Burke GL, Savage PJ, Sidney S, Gardin JM, Oberman A. Exercise Blood Pressure Response and 5-Year Risk of Elevated Blood Pressure in a Cohort of Young Adults: The CARDIA Study. American Journal of Hypertension. 1994;7:234–241.

8. Landsteiner I, Stolze LK, Peterson TE, et al. Multiorgan Physiological Deficits During Exercise Identify Clinical and Molecular Predisposition to Heart Failure With Preserved Ejection Fraction. Circulation. 2026;153:1362–1384.

9. Nayor M, Gajjar P, Murthy VL, et al. Blood Pressure Responses During Exercise: Physiological Correlates and Clinical Implications. *Arteriosclerosis*, Thrombosis, and Vascular Biology. 2023;43:163–173.

10. Cuspidi C, Faggiano A, Gherbesi E, Sala C, Grassi G, Tadic M. Clinical and Prognostic Value of Exaggerated Blood Pressure Response to Exercise. Rev Cardiovasc Med. 2023;24:64.

11. Lee D, Artero EG, Sui X, Blair SN. Review: Mortality trends in the general population: the importance of cardiorespiratory fitness. J Psychopharmacol. 2010;24:27–35.

12. Schultz MG, Otahal P, Picone DS, Sharman JE. Clinical Relevance of Exaggerated Exercise Blood Pressure. Journal of the American College of Cardiology. 2015;66:1843–1845.

13. Wielemborek-Musial K, Szmigielska K, Leszczynska J, Jegier A. Blood Pressure Response to Submaximal Exercise Test in Adults. BioMed Research International. 2016;2016:e5607507.

14. Miyai N, Arita M, Miyashita K, Morioka I, Shiraishi T, Nishio I. Blood Pressure Response to Heart Rate During Exercise Test and Risk of Future Hypertension. Hypertension. 2002;39:761–766.

15. Lewis GD, Gona P, Larson MG, et al. Exercise Blood Pressure and the Risk of Incident Cardiovascular Disease (from the Framingham Heart Study). The American Journal of Cardiology. 2008;101:1614–1620.

16. Miyai N, Shiozaki M, Terada K, et al. Exaggerated blood pressure response to exercise is associated with subclinical vascular impairment in healthy normotensive individuals. Clinical and Experimental Hypertension. 2021;43:56–62.

17. Baycan ÖF, Çelik FB, Güvenç TS, et al. Coronary flow velocity reserve is reduced in patients with an exaggerated blood pressure response to exercise. Hypertens Res. 2022;45:1653–1663.

18. Mariano IM, Amaral AL, Ribeiro PAB, Puga GM. Exercise training improves blood pressure reactivity to stress: a systematic review and meta-analysis. Sci Rep. 2023;13:10962.

19. Mert KU, Şener E, Yılmaz AS, et al. The association of exaggerated hypertensive response to exercise and beta-blockers use in hypertensives. Clinical and Experimental Hypertension. 2020;42:707–713.

20. Lang RM, Badano LP, Mor-Avi V, et al. Recommendations for Cardiac Chamber Quantification by Echocardiography in Adults: An Update from the American Society of Echocardiography and the European Association of Cardiovascular Imaging. Eur Heart J Cardiovasc Imaging. 2015;16:233–271.

21. Schultz MG, La Gerche A, Sharman JE. Cardiorespiratory Fitness, Workload, and the Blood Pressure Response to Exercise Testing. Exercise and Sport Sciences Reviews. 2022;50:25.

22. Steppan J, Barodka V, Berkowitz DE, Nyhan D. Vascular Stiffness and Increased Pulse Pressure in the Aging Cardiovascular System. Cardiology Research and Practice. 2011;2011:263585.

23. Agarwal N, St John J, Van Iterson EH, Laffin LJ. Association of pulse pressure with death, myocardial infarction, and stroke among cardiovascular outcome trial participants. Am J Prev Cardiol. 2024;17:100623.

24. Fletcher GF, Balady GJ, Amsterdam EA, et al. Exercise Standards for Testing and Training. Circulation. 2001;104:1694–1740.

25. Kohli P, Gulati M. Exercise Stress Testing in Women. Circulation. 2010;122:2570–2580.

26. Lang JJ, Prince SA, Merucci K, et al. Cardiorespiratory fitness is a strong and consistent predictor of morbidity and mortality among adults: an overview of meta-analyses representing over 20.9 million observations from 199 unique cohort studies. British Journal of Sports Medicine. 2024;58:556–566.

27. Kaminsky LA, Myers J, Brubaker PH, et al. 2023 update: The importance of cardiorespiratory fitness in the United States. Progress in Cardiovascular Diseases. 2024;83:3–9.

28. Kokkinos P, Faselis C, Samuel IBH, et al. Cardiorespiratory Fitness and Mortality Risk Across the Spectra of Age, Race, and Sex. Journal of the American College of Cardiology. 2022;80:598–609.

29. Lindley KJ, Barker C, Mahmoud Z, et al. Decreased postpartum exercise capacity after a diagnosis of pre-eclampsia: Implications for CVD risk prediction. American Heart Journal. 2024;275:192–199.

30. Lane-Cordova AD, Carnethon MR, Catov JM, et al. Cardiorespiratory fitness, exercise haemodynamics and birth outcomes: the Coronary Artery Risk Development in Young Adults Study. BJOG. 2018;125:1127–1134.

31. Sarzynski MA, Rice TK, Després J-P, et al. The HERITAGE Family Study: A Review of the Effects of Exercise Training on Cardiometabolic Health, with Insights into Molecular Transducers. Med Sci Sports Exerc. 2022;54:S1–S43.

32. Davis ME, Blake C, Perrotta C, Cunningham C, O’Donoghue G. Impact of training modes on fitness and body composition in women with obesity: A systematic review and meta-analysis. Obesity. 2022;30:300–319.

33. Scholten RR, Thijssen DJH, Lotgering FK, Hopman MTE, Spaanderman MEA. Cardiovascular effects of aerobic exercise training in formerly preeclamptic women and healthy parous control subjects. American Journal of Obstetrics and Gynecology. 2014;211:516.e1–516.e11.

34. McLean MK, Kozai AC, Lane AD. Activity Behaviors: Influencers of Cardiometabolic Risk and Adverse Pregnancy Outcomes. Exercise and Sport Sciences Reviews. 2025;53:178.

35. Countouris ME, Koczo A, Reynolds HR, et al. Characteristics of Premature Myocardial Infarction Among Women With Prior Adverse Pregnancy Outcomes. JACC: Advances. 2023;2:100411.

36. Countouris ME, Catov JM, Zhu J, et al. Association of Hypertensive Disorders of Pregnancy With Coronary Microvascular Dysfunction 8 to 10 Years After Delivery. Circulation: Cardiovascular Imaging. 2024;17:e016561.

37. Ghossein-Doha C, Thilaganathan B, Vaught AJ, Briller JE, Roos-Hesselink JW. Hypertensive Pregnancy Disorder, an Under-Recognized Women Specific Risk Factor for Heart Failure? Eur J Heart Fail. 2025;27:459–472.

38. Williams D, Stout MJ, Rosenbloom JI, et al. Preeclampsia Predicts Risk of Hospitalization for Heart Failure With Preserved Ejection Fraction. JACC. 2021;78:2281–2290.

39. Countouris ME, Bello NA. Advances in Our Understanding of Cardiovascular Diseases After Preeclampsia. Circulation Research. 2025;136:583–593.

